# Sex-specific differences in COVID-19 testing, cases and outcomes: a population-wide study in Ontario, Canada

**DOI:** 10.1101/2020.04.30.20086975

**Authors:** Nathan M. Stall, Wei Wu, Lauren Lapointe-Shaw, David N. Fisman, Michael P. Hillmer, Paula A. Rochon

## Abstract

In this population-wide study in Ontario, Canada we report on all 194,372 unique residents who received testing for SARS-CoV-2 between January 23, 2020 and April 28, 2020. We found that while more women than men were tested for SARS-CoV-2, men had a higher rate of laboratory-confirmed COVID-19 infection, hospitalization, ICU admission and death. These findings were consistent even with age adjustment, suggesting that the observed differences in outcomes between women and men were not explained by age or systematic differences in testing by sex. Instead, they may be due to sex-based immunological or other gendered differences, such as higher rates of smoking leading to cardiovascular disease.

## Introduction

The Severe Acute Respiratory Syndrome Coronavirus 2 (SARS-CoV-2) emerged in Wuhan, China in late 2019 and spread globally resulting in the COVID-19 pandemic.^1^ During the two previous coronavirus epidemics, Severe Acute Respiratory Syndrome (SARS) and Middle East Respiratory Syndrome (MERS), male sex was associated with worse clinical outcomes.^2^ Though there is limited sex-disaggregated COVID-19 incidence and outcome data, reports from epidemics in Italy and China indicate that men may also be more affected.^1,3^ It is unclear whether these findings may be skewed because of unreported sex-based differences in SARS-CoV-2 testing and the age distributions of study populations.^4^

## Methods

This population-wide cohort study included all residents of Ontario, Canada who received a nasopharyngeal swab for SARS-CoV-2 between January 23, 2020 (date swab was performed for first reported case of COVID-19 in Canada) and April 28, 2020. We excluded individuals with unknown sex. Ontario is Canada’s most populous province and is home to nearly 15 million residents who receive universal access to medically necessary services including laboratory testing for SARS-CoV-2 under a publicly funded provincial health insurance program. We obtained data for this study from the Ontario Ministry of Health as part of the province’s emergency “modeling table”, including deidentified line level data on all SARS-CoV-2 testing via the Ontario Laboratories Information System (OLIS) and from the integrated Public Health Information System (iPHIS) for all reported COVID-19 cases and related clinical outcomes.

We reported sex- and age-disaggregated data on SARS-CoV-2 testing, COVID-19 cases and related rates of hospitalization, intensive care unit (ICU) admission and death. We used census data from Statistics Canada to compare sex-based testing by age with the sex and age distribution of the Ontario population. Among laboratory confirmed COVID-19 cases, we used logistic regression to estimate sex-based odds ratios for hospitalization, ICU admission and death, adjusting for 10-year age intervals, with the level of statistical significance set at α = .05. All analyses were performed using SAS statistical software, version 9.4 (SAS Institute Inc.). The study was approved by the Research Ethics Board of the University of Toronto.

## Results

A total of 194,372 unique Ontario residents (122,205 women [62.9%] vs. 72,167 men [37.1%]) received testing for SARS-CoV-2 between January 23, 2020 and April 28, 2020 (Table 1). There were 141 individuals excluded because of unknown sex. With the exception of two age groups (ages 0-9 years and 70-79 years), men received less testing for SARS-CoV-2 than would be expected for their age-based representation in the Ontario population (Table 1).

**Table 1:** Sex- and age-disaggregated SARS-CoV-2 testing and positivity in Ontario, Canada (January 23-April 28, 2020)

| Age (years) | SARS-CoV-2 Testing (n = 194,372) |  | Percentage of men in Ontario (2019) * | Test Positivity (n = 15,587) |  |
| --- | --- | --- | --- | --- | --- |
|  | Women | Men |  | Women | Men |
| All ages, n (%) | 122,202 (62.9%) | 72,167 (37.1%) | 49.4% | 9,041 (7.4%) | 6,546 (9.1%) |
| 0-9, n (%) | 1,895 (45.2%) | 2,298 (54.8%) | 51.1% | 41 (2.2%) | 53 (2.3%) |
| 10-19, n (%) | 2,257 (56.5%) | 1,741 (43.5%) | 51.1% | 146 (6.5%) | 122 (7.0%) |
| 20-29, n (%) | 14,682 (66.0%) | 7,554 (34.0%) | 51.9% | 998 (6.8%) | 727 (9.6%) |
| 30-39, n (%) | 19,432 (65.4%) | 10,277 (34.6%) | 49.9% | 1,048 (5.4%) | 798 (7.8%) |
| 40-49, n (%) | 19,820 (68.2%) | 9,244 (31.8%) | 48.7% | 1,213 (6.1%) | 885 (9.6%) |
| 50-59, n (%) | 21,175 (65.2%) | 11,307 (34.8%) | 49.6% | 1,513 (7.1%) | 1,094 (9.7%) |
| 60-69, n (%) | 13,829 (56.9%) | 10,479 (43.1%) | 48.3% | 1,034 (7.5%) | 1,003 (9.6%) |
| 70-79, n (%) | 9,037 (51.4%) | 8,555 (48.6%) | 46.6% | 699 (7.7%) | 724 (8.5%) |
| ≥80, n (%) | 20,078 (65.2%) | 10,712 (34.8%) | 40.0% | 2,349 (11.7%) | 1,140 (10.6%) |

\*Census-data from Statistics Canada (Statistics Canada. Table 17-10-0005-01 Population estimates on July 1st, by age and sex. https://www150.statcan.gc.ca/t1/tbl1/en/cv.action?pid=1710000501).

Compared to women, men had a higher rate of laboratory-confirmed COVID-19 infection (6,546 men [9.1%] vs. 9,041 women [7.4%]), a finding consistent across all age groups (Table 1). Among all individuals with COVID-19 infection, men had higher rates of hospitalization (998 men [15.3%] vs. 798 women [8.9%]), ICU admission (294 men [4.5%] vs. 154 women [1.7%]), and death (477 men [7.3%] vs. 509 women [5.6%]) (Table 2). In age-adjusted analyses, male sex was associated with a higher odds of hospitalization (adjusted odds ratio [aOR], 1.82, 95% confidence interval [CI], 1.64-2.01, P<0.001), ICU admission (aOR, 2.39, 95% CI, 1.96-2.92, P<0.001), and death (aOR, 1.75, 95% CI, 1.52-2.01, P<0.001) (Table 2).

**Table 2:** Sex- and age-disaggregated COVID-19 clinical outcomes in Ontario, Canada (January 23-April 28, 2020)

|  | Women | Men | aOR (95% CI)<br>Men vs. Women | P-Value |
| --- | --- | --- | --- | --- |
| **All ages** | n = 9,041 | n = 6,546 |  |  |
| Hospitalization | 798 (8.9%) | 998 (15.3%) | 1.82 (1.64-2.01) | <0.001 |
| ICU admission | 154 (1.7%) | 294 (4.5%) | 2.39 (1.96-2.92) | <0.001 |
| Death | 509 (5.6%) | 477 (7.3%) | 1.75 (1.52-2.01) | <0.001 |
| **Age 0-59 years** | n = 4,959 | n = 3,679 |  |  |
| Hospitalization | 251 (5.1%) | 359 (9.8%) | 2.09 (1.76-2.47) | <0.001 |
| ICU admission | 54 (1.1%) | 122 (3.3%) | 3.21 (2.32-4.44) | <0.001 |
| Death | 19 (0.4%) | 33 (0.9%) | 2.41 (1.37-4.24) | 0.002 |
| **Age 60-79 years** | n = 1,733 | n = 1,727 |  |  |
| Hospitalization | 181 (17.0%) | 414 (24.0%) | 1.54 (1.30-1.82) | <0.001 |
| ICU admission | 75 (4.3%) | 152 (8.8%) | 2.13 (1.60-2.83) | <0.001 |
| Death | 97 (5.6%) | 166 (9.6%) | 1.79 (1.37-2.32) | <0.001 |
| **Age ≥80 years** | n = 2,349 | n = 1,140 |  |  |
| Hospitalization | 253 (10.8%) | 225 (19.7%) | 1.89 (1.55-2.30) | <0.001 |
| ICU admission | 25 (1.1%) | 20 (1.8%) | 1.39 (0.76-2.52) | 0.28 |
| Death | 393 (16.7%) | 278 (24.4%) | 1.68 (1.41-2.00) | <0.001 |

aOR = adjusted odds ratio; CI = confidence interval; ICU = intensive care unit

## Discussion

We found that while more women than men were tested for SARS-CoV-2, men had a higher rate of laboratory-confirmed COVID-19 infection, hospitalization, ICU admission and death. These findings were consistent even with age adjustment, suggesting that the observed differences in outcomes between women and men were not explained by age or systematic differences in testing by sex. Instead, they may be due to sex-based immunological or other gendered differences, such as higher rates of smoking leading to cardiovascular disease.^5^

The study is limited to a single region and could not control for underlying differences in sociodemographic characteristics and comorbidities between women and men. We also could not identify healthcare workers—the majority of whom are women—which could explain some of the sex-based differences in SARS-CoV-2 testing. With the majority of regional health systems failing to report fully sex-disaggregated data on COVID-19, our study highlights how sex-specific reporting can guide a more gender-responsive approach to the global pandemic.^2,5^ In particular, our findings can inform pathways for COVID-19 care, including targeting older men as a particularly at risk group who may benefit from intensified prevention and earlier intervention.^2^

## Data Availability

The underlying analytic code are available from the authors upon request (e-mail), understanding that the computer programs may rely upon coding templates or macros that are unique and are therefore either inaccessible or may require modification.

## References

1. The Novel Coronavirus Pneumonia Emergency Response Epidemiology Team. Vital Surveillances: The Epidemiological Characteristics of an Outbreak of 2019 Novel Coronavirus Diseases (COVID-19) — China, 2020. China CDC Weekly Web site. http://weekly.chinacdc.cn/en/article/doi/10.46234/ccdcw2020.032. Published 2020. Accessed April 30, 2020.

2. Purdie A, Hawkes S, Buse K, et al. Sex, gender and COVID-19: Disaggregated data and health disparities. BMJ Global Health Blog Web site. https://blogs.bmj.com/bmjgh/2020/03/24/sex-gender-and-covid-19-disaggregated-data-and-health-disparities/. Published 2020. Updated March 24, 2020. Accessed April 19, 2020.

3. Istituto Superiore di Sanità. Characteristics of SARS-CoV-2 patients dying in Italy Report based on available data on April 23th, 2020. https://www.epicentro.iss.it/en/coronavirus/bollettino/Report-COVID-2019_23_april_2020.pdf. Published 2020. Accessed April 30, 2020.

4. Global Health 5050. COVID-19 sex-disaggregated data tracker. http://globalhealth5050.org/Covid19/. Published 2020. Accessed April 19, 2020.

5. Wenham C, Smith J, Morgan R, Gender, Group C-W. COVID-19: the gendered impacts of the outbreak. Lancet. 2020;395(10227):846–848.

